# RPA-Based Method For The Detection Of SARS-COV2

**DOI:** 10.1101/2020.09.17.20196402

**Authors:** Angus A. Nassir, Mazarati Jean Baptiste, Ivan Mwikarago, Majidi R. Habimana, Janvier Ndinkabandi, Anthere Murangwa, Thierry Nyatanyi, Claude Mambo Muvunyi, Sabin Nsanzimana, Mutesa Leon, Clarisse Musanabaganwa

**Affiliations:** Bioinformatics Institute of Kenya, Nairobi, Kenya; Medical Research Center, Rwanda Biomedical Center, Kigali, Rwanda; National Reference Laboratory, Rwanda Biomedical Center, Kigali, Rwanda; National COVID19 Taskforce, Kigali, Rwanda; University of Rwanda, Kigali, Rwanda; Rwanda Military Hospital, Kigali, Rwanda

**Keywords:** RPA, COVID-19

## Abstract

**Background:** Coronavirus disease 2019 (COVID-19) is a highly infectious disease with significant mortality, morbidity, and far-reaching economic and social disruptions. Testing is key in the fight against COVID-19 disease. The gold standard for COVID-19 testing is the reverse transcription polymerase chain reaction (RT-PCR) test. RT-PCR requires highly specialized, expensive, and advanced bulky equipment that is difficult to use in the field or in a point of care setting. There is need for a simpler, inexpensive, convenient, portable and accurate test. Our aims were to: (i) design primer-probe pairs for use in isothermal amplification of the S1, ORF3 and ORF8 regions of the SARS-CoV2 virus; (ii) optimize the recombinase polymerase amplification (RPA) assay for the isothermal amplification of the named SARS-COV2 regions; (iii) detect amplification products on a lateral flow device. and (ii) perform a pilot field validation of RPA on RNA extracted from nasopharyngeal swabs.

**Results:** Assay validation was done at the National Reference Lab (NRL) at the Rwanda Biomedical Center (RBC) in Rwanda. Results were compared to an established, WHO-approved rRT-PCR laboratory protocol. The assay provides a faster and cheaper alternative to rRT-PCR with 100% sensitivity, 93% specificity, and positive and negative predictive agreements of 100% and 93% respectively.

**Conclusion:** To the best of our knowledge, this is the first in-field and comparative laboratory validation of RPA for COVID-19 disease in low resource settings. Further standardization will be required for deployment of the RPA assay in field settings.

## Background

Coronavirus disease 2019 (COVID-19) is a highly infectious disease that has rapidly spread all over the world leading to thousands of deaths, hospitalizations, and far-reaching economic and social disruptions (Guo *et al*., 2020). COVID-19 disease is caused by the severe acute respiratory syndrome coronavirus 2 (SARS-CoV-2) and was first reported in Wuhan, China, in December, 2019 (Wu *et al*., 2020). Fever and cough are the most common symptoms of the infection. other symptoms include fatigue, dyspnea, muscle ache, headache, chest pain, diarrhea, hemoptysis, sputum production, rhinorrhea, nausea and vomiting, sore throat, confusion, diarrhea, and anorexia (Wu & McGoogan, 2020). The virus has caused more than 24 million infections and 824,000 deaths worldwide with massive social disruption and far-reaching economic effects as at the end of August 2020 (WHO, 2020).

Transmission of SARS-CoV2 is from person to person through respiratory droplets due to coughing and sneezing and airborne aerosols. The disease can also spread through touching infected surfaces and via the fecal-oral route (Lotfi *et al*., 2020). Clinical signs of COVID-19 disease appear 2-14 days after exposure to the virus and include fever, sore throat, cough, headache, shortness of breath or difficulty breathing, muscle pain, chills, repeated shaking with chills, and new loss of taste or smell (Chen *et al*., 2020; Sahin *et al*., 2020).

Testing is key in the fight against COVID-19 disease and is vital for the reopening of economies. The gold standard for COVID-19 testing is the real-time reverse transcription polymerase chain reaction (RT-PCR) (WHO, 2020). Whereas RT-PCR produces accurate and reproducible results, it requires highly specialized, expensive, and advanced bulky equipment which cannot be used in the field or in a point of care setting. It is also slow, and requires highly trained scientists. Additionally, current RT-PCR tests are based on invasive sampling since samples are collected using nasopharyngeal swabs which may be uncomfortable to many (Tang *et al*., 2020).

The antigen/antibody tests for COVID-19 overcome many of the challenges of the RT-PCR tests since they are faster to conduct, cheaper, do not require expensive and bulky equipment, and can be conducted in the field or at a point of care setting by non-scientists. However, the antigen antibody tests are not very reliable and have a lower accuracy rate (Deeks *et al*., 2020). Therefore, there is need for a test that provides the convenience and ease of the antigen/ antibody test but with the accuracy and reliability of the RT-PCR test.

We have optimized an RPA assay for the detection of SARS-CoV2 for use in the field and in rural settings and tested its applicability at a field station in Kigali, Rwanda. The assay leverages on the power of recombinase polymerase amplification and lateral flow detection to provide a method for the rapid detection of the SARS CoV2 in an accurate, simple, and affordable manner. Unlike the RT-PCR test, isothermal amplification with detection on a lateral flow device does not require expensive equipment, is portable and easy to perform, and can be done in the field or in a point of care setting. It does not require highly trained scientists, is portable, versatile, and does not require stable electricity (Daher *et al*., 2016). The optimized RPA assay was compared against the LightMix RT-PCR test (Roche, Basel, Switzerland) as the ‘gold standard’.

## Methods

### Design of Primers and Probes

Primer design was based on the complete genome sequence of the SARS CoV2 isolate Wuhan-Hu-1, complete genome, NCBI Reference Sequence: NC_045512.2 (Appendix 1). The SARS CoV 2 genome is 29891 nucleotides in size, and it encodes 9860 amino acids with a G + C content of 38%. Structurally, the genome is composed of two flanking untranslated regions (UTRs) and one long open reading frame encoding a polyprotein arranged as follows orf1a-orf1b-orf2-orf10. The 5’ UTR region is 265 nucleotides long while the 3’ UTR region is 358 nucleotides long. The Orf1/ab encodes 5′-replicase (orf1/ab)-structural proteins namely the Spike (S), Envelope (E), Membrane (M), and Nucleocapsid (N) proteins. The replicase proteins are cleaved into 16 non-structural proteins (nsps) (Khailany *et al*., 2020).

To determine the best regions to target for efficient primer design, BLAST analysis was performed and the percent sequence identity between the SARS CoV2 sequences and other coronavirus sequences determined (table S 1). Primers were designed using Primer3 Plus software to flank the Orf3b and Orf8/8b regions since this exhibit the lowest homology between SARS CoV2 and other coronaviruses (Untergasser *et al*., 2012). This helped maximize the specificity of the test assay. Additional primers targeted at the S1 region were designed (figure 1-9).

**Figure 1:**
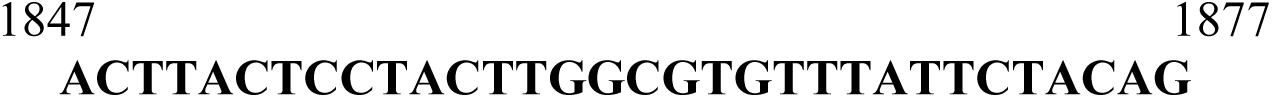
Forward primer for the SARS-CoV2 S1 region.

The rationale behind the design of S1 primers is that the S precursor protein of SARS-CoV-2 can be proteolytically cleaved into S1 (685 aa) and S2 (588 aa) subunits. The S1 subunit contains a signal peptide, followed by an N-terminal domain (NTD) and receptor-binding domain (RBD), while the S2 subunit contains conserved fusion peptide (FP), heptad repeat (HR) 1 and 2, transmembrane domain (TM), and cytoplasmic domain (CP). The S2 protein is well conserved among SARS-CoV-2 viruses and shares 99% identity with that of bat SARS-CoVs. In contrast, the S1 subunit consists of the receptor-binding domain (RBD), which mediates virus entry into sensitive cells through the host angiotensin-converting enzyme 2 (ACE2) receptor. The S1 protein of 2019-nCoV shares about 70% identity with that of human SARS-CoVs. The highest number of variations of amino acids in the RBD is located in the external subdomain (receptor binding motif, RBM), which is responsible for the direct interaction between virus and host receptor (Tai *et al*., 2020). Our primer target was the more conserved section of the RBD.

Primer design was based on best practice with regard to GC content and clamps and other basic requirements but optimized for length as is needed for RPA probes. Length optimization ensured that all primers lengths were between 18-35 with amplicon sizes ranging from 80-400bp. All forward primers relating to have unmodified 5’ ends and all reverse primers have a biotin moiety conjoined at their 5’ ends (Li *et al., 2019*). The primer pairs amplify S1 region of SARS-CoV2.

ORF3 and orf8a/b probes consisted of a 5’ end labeled with fluorescein amidites (FAM), an internal abasic THF residue, and a 3’ spacer region and is homologous to a region of the amplified ORF3a/ORF8a/b sequence.

### RNA Extraction Procedure

RNA was extracted from nasopharyngeal swabs using the QIAamp® Viral RNA kit (Qiagen, Germany) according to the manufacturer’s instructions. Briefly, 560 μl of lysis buffer + carrier RNA was added into a sterile 1.5 ml centrifuge tube. Into this tube, 140 μl of the sample was added and the mixture vortexed for 15 seconds, then incubated at room temperature for 10 minutes. 560 μl of ethanol was added followed by vortexing for 15 seconds. The whole mixture was drawn into a spin column and centrifugation conducted at 8000rpm for 1 minute at room temperature. The spin column was fit into a new collection tube, 500 μl wash buffer (AW1) was added into the spin column, and centrifugation done at 8000rpm for 1 minute at room temperature. The spin column was fit in a new collection tube and 500 μl wash buffer (AW2) was added into the spin column followed by centrifugation at 14,000rpm and centrifugation for 3 minutes at room temperature. The spin column and collection tube was spun at 14,000rpm centrifugation for 1 minutes to remove residual ethanol. RNA was eluted using 60 μl eluent and the nucleic acid solution used immediately or stored at -20^0^c for later use. The concentration of RNA was measured using the NanoDrop(tm) 2000/2000c Spectrophotometer (ThermoFisher, UK).

### RT-PCR

RT-PCR was conducted on Applied Biosystems 7500 Fast Real Time PCR System (Roche, Basel, Switzerland) using the LightMix RT-PCR test kit (Roche, Basel, Switzerland). The thermal cycling condition was 45 °C for 10 min, 95 °C for 10 min, followed by 45 cycles of 95 °C for 15 s, and 55 °C for 1 minute. Negative controls (no template) were incorporated into each set of reactions. Cycle threshold (ct) values higher than 40 were considered to be negative results.

### RPA Optimization

The TwistAmp nfo Kit (TANFO02KIT; TwistDX, Maidenhead, UK) was used to perform the reactions in a final volume of 48 μl. The reaction mix was prepared in a 1.5 ml tube. The master mix comprised of 2.1 μl of 10 μM forward primer, 2.1 μl of 10. 2 μl M reverse primer, TwistAmp® nfo probe (0.6ul), 27.5 μl of the rehydration buffer, 2.5 μl of SuperScript(tm) III Reverse Transcriptase (ThermoFisher, UK), and 2 μl of RNase H (ThermoFisher, UK). RNA template (1ul) and water up to 10 μl was added to the mastermix and thorough mixing done. The reaction mix was added to a TwistAmp® nfo reaction and mixed by pipetting. 2.5 μl of 280mM Magnesium Acetate (MgOAc) was added to the reaction and the reaction started by mixing. Reactions were incubated in a heat block at 37°C for 20 minutes and after 4 minutes the strips were removed, vortexed, and spun briefly, then returned into the heating block. Negative controls (no template) were incorporated into each set of reactions.

### Detection on a Lateral Flow Device

The FAM-sequence-biotin amplicons were detected on a PCRD lateral flow cassette (Abingdon Health, York, UK) where presence of the viral material was indicated by formation of a line on the 2^nd^ position of the cassette. To detect the amplicons, 6 μl of the amplification product was pipetted into a 1.5 ml tube and diluted using 84 μl of the PCRD extraction buffer. The foiled device was opened immediately before use and 75 μl of the diluted reaction mixture added to the sample well of the PCRD cassette. The cassette was left in the horizontal position for 10 minutes and then read visually. The third position of the PCRD lateral flow cassette has a control line that forms to show that fluid has passed in the flow cell. Positive samples therefore had 2 lines forming on the cassette: line 1 positive for S1 and line 3 which is the control line.

### Validation Assays at NRL, RBC

Assay validation was done at the National Reference Lab (NRL) at the Rwanda Biomedical Center (RBC). Previously collected, anonymized samples from current COVID-19 patients were randomly selected and used to validate the assay. The cycle threshold of the patient samples used ranged between 16-39 (table S 2). The template average concentration was calculated using the following formula N1 V1=N2 V2 by considering the standard concentration of 28.9 ng/ μl for sample B32213 with ct Values ranging 36-37 (Figure S 1). The sensitivity and specificity of the assay was calculated and the measures validated by RT-PCR using the LightMix®Modular SARS-CoV2 E-gene and RdRp kit (Roche, Basel, Switzerland) according to WHO and Rwanda NRL guidelines. Visualization was done using the PCRD cassettes

### Statistical Tests

Graphpad QuickCalcs (https://www.graphpad.com/quickcalcs/ConfInterval1.cfm) was used to calculate confidence intervals (95% CI) for sensitivity and specificity comparisons of LightMix®Modular SARS-CoV2 RT-PCR and RPA. The easyRoc v1.3.1 (http://www.biosoft.hacettepe.edu.tr/easyROC/) tool was used to calculate receiver operating characteristic (ROC) curves and the sensitivity, specificity, and negative and positive predictive values and likelihood ratios for comparison of the RPA assay against the LightMix®Modular SARS-CoV2 RT-PCR as the ‘gold standard.’ *P*-values of < 0.05 were considered significant. SE estimation was done using the DeLong (1988) method and the Youden index used for the optimal cut-off (Goksuluk *et al*., 2016).

## Results

Of the 28 samples that were tested using the LightMix®Modular SARS-CoV2 E and RdRp gene kits, 50% (n=14) were positive and 50% (n=14) were negative. All these samples were subjected to RPA analysis. Of the 14 that were positive by LightMix®Modular SARS-CoV2 RT-PCR, 13 (∼93%) were positive by RPA. All the 14 samples that were negative by LightMix®Modular SARS-CoV2 RT-PCR were also negative by RPA (100%) (Table 1).

**Table 1:** Results of samples assayed by LightMix®Modular SARS-CoV2 E and RdRp gene RT-PCR test and RPA assay (n = 28)

|  | Positive | Negative |
| --- | --- | --- |
| **LightMix®Modular SARS-CoV2 E-gene** | 14 | 14 |
| **LightMix®Modular SARS-CoV2 RdRp gene** | 14 | 14 |
| **RPA Assay** | 13 | 14 |
| **Positive % agreement** | 93% |  |
| **Negative % agreement** |  | 100% |

Samples positive for the SARS-CoV2 S1 gene had 2 lines while those negative for the SARS-CoV2 S1 gene had only a single control line. All samples were retested for reproducibility and the same results were obtained, giving 100% reproducibility (figure 1).

Cutoff values for the RPA assay including the specificity, sensitivity, PPV, NPV, NLR, and PLR are shown in table 1.

**Table 1:**
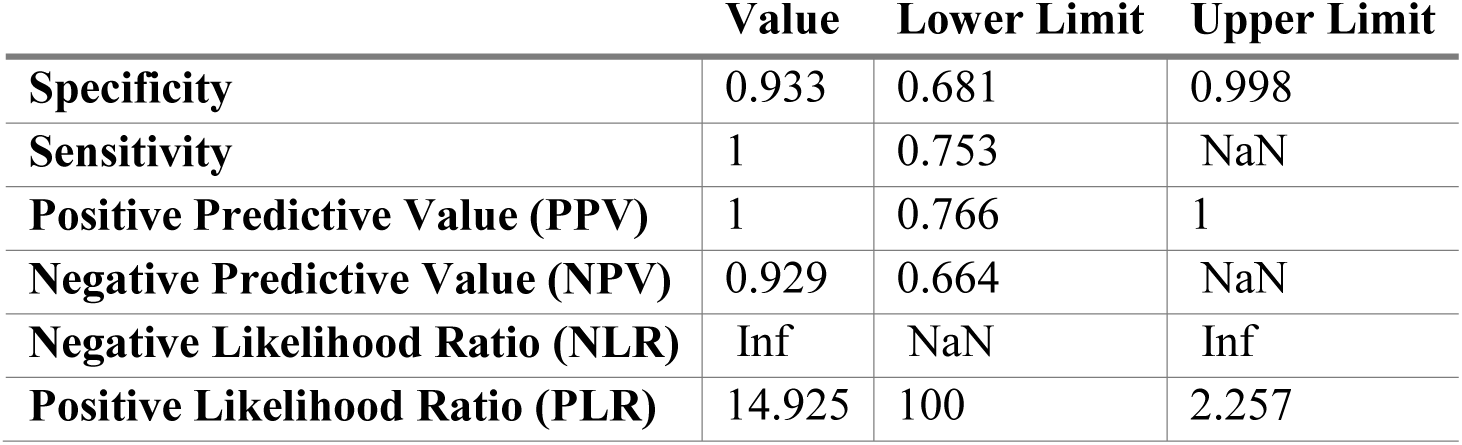
Cutoff Results for the RPA assay.

|  | Value | Lower Limit | Upper Limit |
| --- | --- | --- | --- |
| **Specificity** | 0.933 | 0.681 | 0.998 |
| **Sensitivity** | 1 | 0.753 | NaN |
| **Positive Predictive Value (PPV)** | 1 | 0.766 | 1 |
| **Negative Predictive Value (NPV)** | 0.929 | 0.664 | NaN |
| **Negative Likelihood Ratio (NLR)** | Inf | NaN | Inf |
| **Positive Likelihood Ratio (PLR)** | 14.925 | 100 | 2.257 |

Figure 2 shows the corresponding ROC curve for the RPA assay. SE and confidence interval (CI) estimation was done using the DeLong method (DeLong *et al*., 1988). For RPA, the area under the curve (AUC) was 0.97 (95% CI: 0.6646 to >0.9999) (P=1.357831e-74). The true positive rate (TPR) for the RPA assay was 92.9% and the false positive rate (FPR) was 0%.

**Figure 2:**
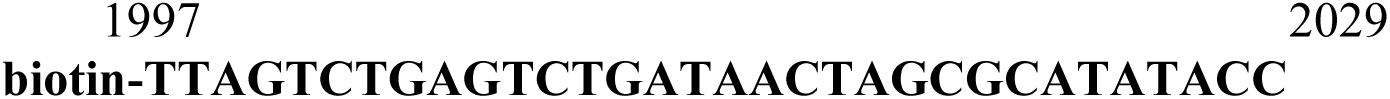
Reverse primer for the SARS-CoV2 S1 region.

**Figure 3:**
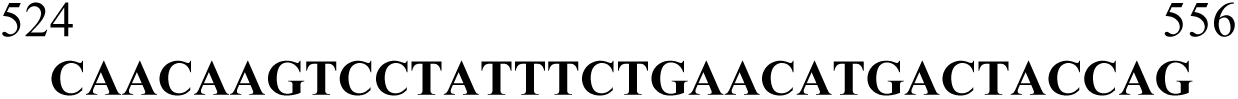
Forward primer for the SARS-CoV2 ORF3a region.

**Figure 4:**
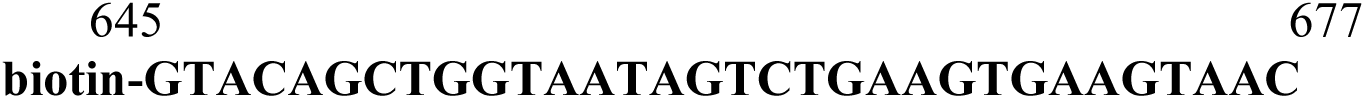
Reverse primer for the SARS-CoV2 ORF3a region.

**Figure 5:**
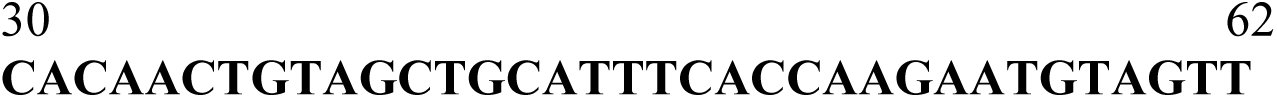
Forward primer for the SARS-CoV2 ORF8 region.

**Figure 6:**
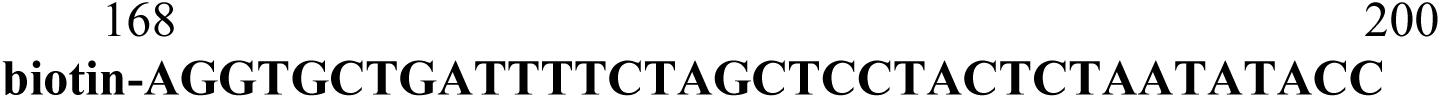
Reverse primer for the SARS-CoV2 ORF8 region.

**Figure 7:**
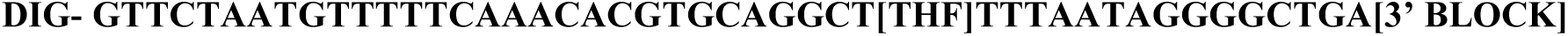
SARS-CoV2 S1 region probe.

**Figure 8:**
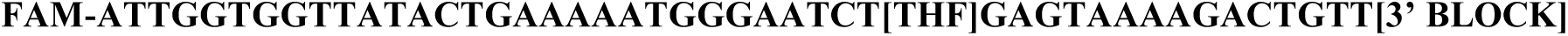
SARS-CoV2 ORF3a region probe.

**Figure 9:**
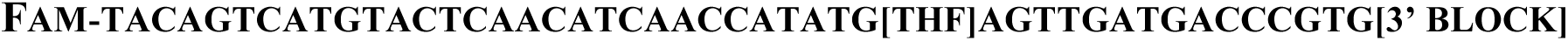
SARS-CoV2 ORF8 region probe.

**Figure 10:**
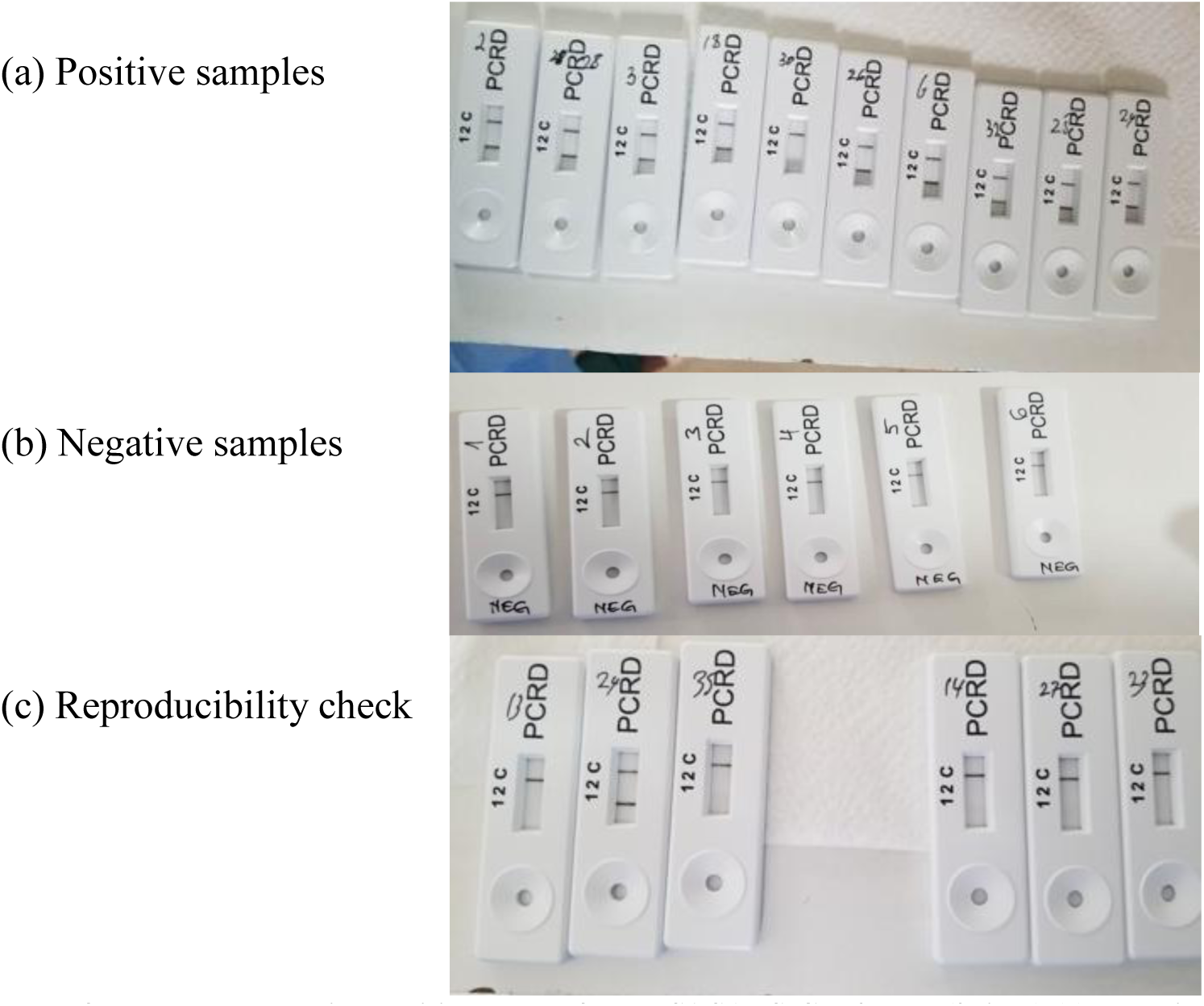
Lateral flow cassettes showing positive results for the S1 SARS-CoV2 gene (2 lines) (a), negative RPA results for the S1 SARS-CoV2 gene (a single control line) (b), and reproducibility check results (c)

**Figure 11:**
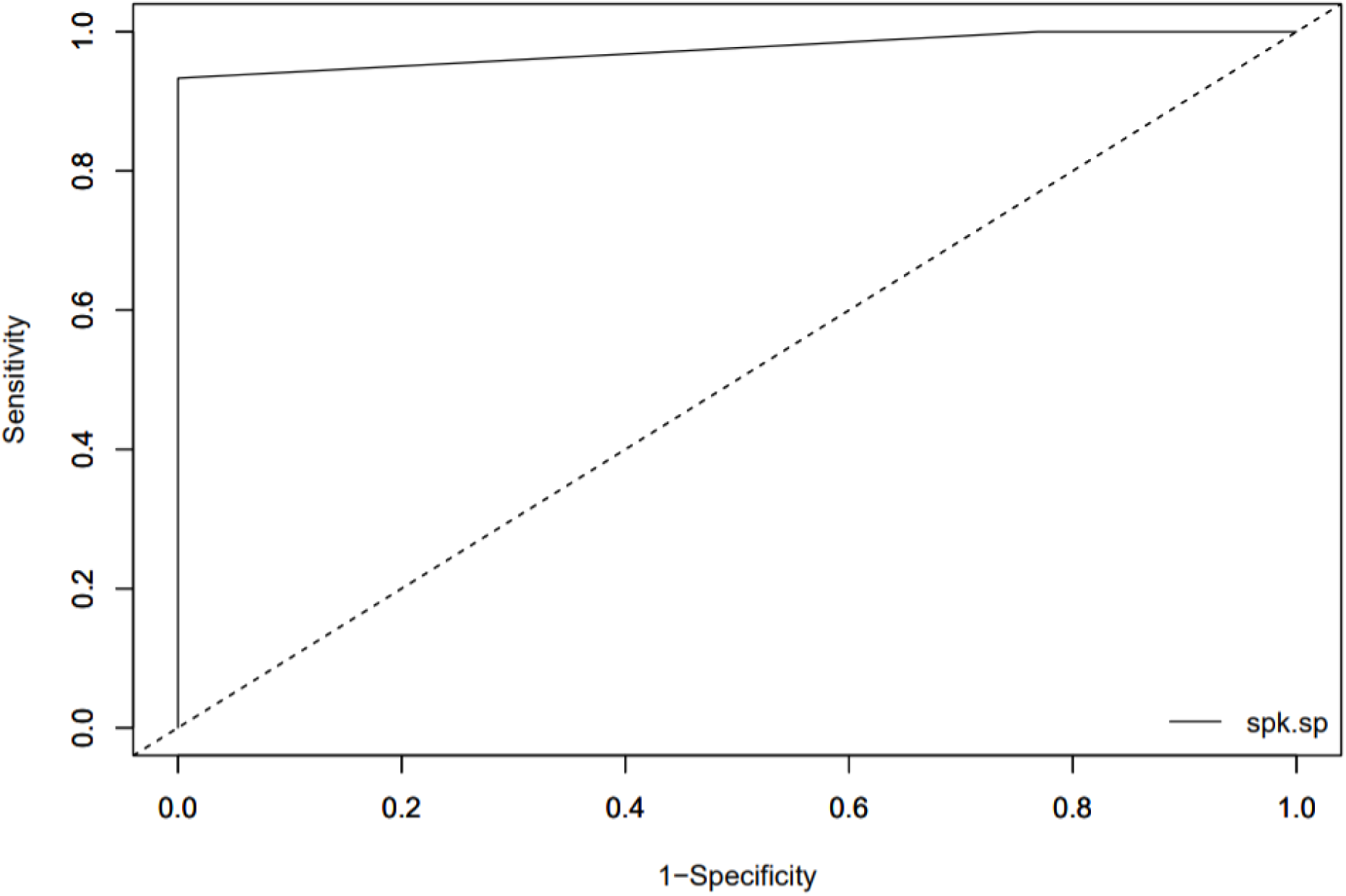
ROC Curves of RPA against LightMix®Modular SARS-CoV2 RT-PCR.

The area under the curve (AUC) was 0.97 (table 2).

**Table 2:** AUC calculations.

| Marker | AUC | SE.AUC | LowerLimit | UpperLimit (*) | z | p-value |
| --- | --- | --- | --- | --- | --- | --- |
| spk.sp | 0.974359 | 0.02596 | 0.923479 | 1.025239 | 18.273 | 1.36E-74 |

The same samples were tested again using a second round of the RPA assay to determine whether the similar results would be obtained. 11/ 13 samples that had tested positive in the first RPA assay turned positive suggesting a reproducibility rate of 84% while one of the samples that had tested positive in the first round assay turned negative for the second time. All the 14 samples that were negative in the first RPA assay also tested negative again, suggesting a reproducibility rate of 100% for negative samples.

## Discussion

In this study, we have shown that the RPA assay has a sensitivity of 100%, and specificity of ∼93% compared to the gold standard RT-PCR test when used in the detection of the SARS-CoV2 virus from nasopharyngeal swabs. The gold standard was the LightMix® Modular SARS-CoV-2 RT-PCR kit targeting the envelope (E) and RNA-dependent RNA polymerase (RdRp) genes. The LightMix® Modular SARS-CoV-2 E-gene and RdRp gene tests are based on amplification of a 76 bp long fragment from a conserved region in the SARS-CoV2 E gene and of the RdRp gene respectively followed by detection using FAM-labelled hydrolysis probes. The sensitivity and specificity of the LightMix® E-gene kit has been validated by several researchers (Corman *et al*., 2020; Roche, 2020). Findings show that this test has high specificity and sensitivity, does not show cross reactions with common pathogens, is rapid with fewer verification requirements, and is widely deployed (Yip *et al*., 2020; PHE, 2020). Based on this, the LightMix® Modular SARS-CoV-2 RT-PCR kit was an appropriate gold standard to use for comparison during the RPA validation.

The overall sensitivity of the RPA assay was 93.3%, specificity 100%, positive predictive value 100%, negative predictive value 92.9%, and the negative likelihood ratio 0.067 compared to the gold standard. A systematic review of RT-PCR tests revealed that initial false-negative RT-PCR results occur in 54% of all COVID-19 tests and that the sensitivity of these tests ranges from 71-98% (Arevalo-Rodriguez *et al*., 2020). Broughton *et al* (2020) described an RPA-based method for detection of SARS-CoV2 using CRISPR/Cas-12 and reported a PPV of 95% and 100% NPV compared to the conventional RT-PCR assay (Broughton *et al*., 2020). Thus, our RPA assay compares well with available RT-PCR tests and the CRISPR/Cas12 test.

The positive likelihood ratio (LR) of our assay was infinite. Infinite LR suggest that the condition that’s being tested is definitely present. On the other hand, negative LR, which is the ratio of true negatives to false negatives, acts as a pointer on the absence of the condition being tested given a negative result (Deeks & Altman, 2004). The negative LR for our RPA assay was 14.9. Based on the LR ratios, the conclusion is that the assay is asymmetric as it is very good at ruling in covid-19 infection when the assay is positive than it is at ruling out infection when the test is negative. Predictive values were 1.0 (positive) and 0.929 (negative) meaning that a positive RPA assay is 100% predictive of SARS-CoV2 infection but a negative RPA assay result leaves a 7% probability of disease.

The ROC curve is a plot of sensitivity versus 1-specificty and provides mean sensitivity values over all possible specificity values. The converse is also true. The ROC is a good measure of a diagnostic test’s utility (Mandrekar, 2010). The AUC in our assay was 0.97. According to Hosmer & Lemeshow (2000), AUC values greater than 0.9 are outstanding. This implies that the RPA assay has excellent discriminating ability and will correctly distinguish covid-19 positive samples correctly from negative samples 97% of the time (Hosmer & Lemeshow, 2000)

Out of the 14 samples that tested positive with the gold standard, only one was negative for the RPA assay. There are several possible explanations for this failure including variation due to use of different gene primers, mutations in the SARS-CoV2 region targeted by the S1 gene primer-probe pairs, personnel skills, technical factors, lab standards, sampling procedures, contamination of the RT-PCR sample, and lack of optimization of enzyme concentrations and assay reaction temperatures. This can be possibly corrected through use of multiple gene targets involving multiplexing of the other unused ORF3 and ORF8 primer-probe pairs with the S1 primer-probe pair, optimization of enzyme concentrations and assay reaction temperatures, and review of the lab procedures for possible anomalies (Tahamtan & Ardebili, 2020). Reproducibility was 100% for samples that had initially tested negative and 84% for those that had initially tested positive discounting possible anomalies in the lab testing process for the RPA assay.

Even though RT-PCR tests such as the gold standard used in this study have received positive reviews (Corman *et al*., 2020), these tests can also detect other sarbecoviruses including bat SARS-related coronaviruses and SARS-CoV and is not only specific for SARS-CoV-2 in spite of the fact that they are designed around well conserved regions (Corman *et al*., 2020). This means that they are primarily suited for initial screening with the requirement of further confirmatory testing. This non-specificity can be attributed to the significant homology that’s shared between the E envelope of SARS-CoV2 and other coronaviruses (Yip *et al*., 2020). In contrast, our assay utilized primer-probe pairs for the S1 region of the spike glycoprotein which is less homologous to the glycoprotein of other coronaviruses. The SARS-CoV2 E gene shares 100% and 95% homology with Chinese horseshoe bat (*Rhinolophus sinicus*) coronavirus (bat-SL-CoVZXC21) and SARS-CoV E genes respectively. Homology between SARS-CoV2 ORF3b, bat-SL-CoVZXC21, and SARS-CoV2 is 32%. Homology between SARS-CoV2 ORF8 and bat-SL-CoVZXC21 and SARS-CoV2 is 94% and 40% respectively while that between SARS-CoV2, bat-SL-CoVZXC21 and SARS-CoV is 70%. This reduced similarity together with relatively longer primers and probes helps to overcome the weakness of the conventional RT-PCR tests with regard to detection of other sarbecoviruses.

Mutations in the S1 and ORF3b/8 regions may affect the ability of our assay to accurately detect SARS-CoV2. Korber *et al* (2020) reported a widespread mutation in the S gene. According to their findings, the D614G mutation involves replacement of glutamic acid by aspartic acid in the 614^th^ position of the S protein (position 23,403 in the Wuhan reference). The D614G mutation is part of the G clade, it is almost always linked to 3 other mutations, and differs from the first Wuhan virus by 4 mutations. It is suggested that the S mutations may be more infectious (Korber *et al*. 2020). A strong recurrent mutation on the S gene at position 21,575 was reported by van Dorp *et al* (2020). In total, van Dorp *et al* (2020) report 199 SARS-CoV2 mutations. 60% of the reported mutations are found in the ORF1ab region and 11% in the nucleocapsid phosphoprotein region. Lowest mutation rates are found in the E (0.5%), ORF10 (1%), and ORF6 (1%) regions. Mutations in the S, ORF3a, and ORF8 comprise approximately 10%, 6%, and 1.5% of all mutations (van Dorp *et al*., 2020).

From the foregoing, there are 3 conclusions. First, the S1 primers in our RPA assays were designed to span positions 1847 to 1877 (forward) and 1997 to 2029 (reverse) of the S gene. These are, to the best of our knowledge, outside the range of S gene mutations so far described. A similar observation is made for our ORF3 and ORF8 primers. Of all the reported mutations, none has affected the region targeted by our primers yet. Secondly, if such mutations on the S gene are more infectious, the additional utility of our RPA test would be to design primers around the variable region and use this to discriminate mild or less infectious disease (wild type) from the more infectious (mutated) since the assay is based on double detection on a lateral device. The utility would be in multiplexing the S primers with primers from a more conserved region, such as E or ORF8. Thirdly, it is argued that SARS-CoV2 mutation rates are relatively slower and that being a highly infectious disease with a low fatality rate, human hosts may acquire widespread immunity and this may impose selection pressure on the virus, driving new mutations (Biswas *et al*., 2020; Das *et al*., 2020). The implication is that, based on emerging genomic information on SARS-CoV2 mutations, tests may need to be readjusted continuously with regard to primer/probe pairs.

Regarding the concentration of the RNA used in the RPA assay, most of the samples had a concentration below 3 ng/ul. This demonstrates that the RPA assay is very powerful and can detect very small concentrations of the SARS-CoV2 virus and compares well with the minimum RNA concentration required for RT-PCR tests.

### Study Limitations

Due to financial limitations, only 49 samples were tested, optimization of reaction temperatures and MgOAc concentrations was not done, ORF3a and ORF8a/b probes were not tested, and the absolute limit of detection of RNA extracted from nasopharyngeal swabs for RPA testing could not be determined.

## Conclusion

We have shown here that RPA can be applied successfully for the detection of SARS-CoV2 virus in nasopharyngeal swabs. However, further optimization of the reaction temperature and MgOAc concentration is required. Other recommendations include determination of detection limit, testing of crude extracts in place of pure extracts, and multiplexing of S1 and ORF primers and probes in one reaction for double detection of virus as the second step of validation. Findings suggest that sputum is more sensitive than nasal swabs for SARS-Cov2 detection (Yang *et al*., 2020). Since the RPA assay requires minimal RNA concentrations, it is suggested that crude sputum samples be trialed in place of nasal/oropharyngeal swabs since the latter also are invasive and uncomfortable and false negatives have been linked to incorrect collection of nasal swabs. Finally, further validation studies using a larger sample size needs to be conducted.

## Data Availability

The authors declare that the data supporting the findings of this study are available within the article and its supplementary information files

## Availability of data and materials

The findings and conclusions made by this article are supported by data and additional files which have been availed here.

## Acknowledgements

We thank the Rwanda Biomedical Center and the Rwanda COVID-19 National Taskforce for their collaboration and provision of patient samples and access to laboratory facilities.

## Funding

The authors received no financial support for the research, authorship, and/or publication of this article.

## Contributions

AAN performed idea conceptualization, protocol development and study implementation; CM performed study conceptualization, protocol development and study implementation, CM, MH and JN performed the lab work and acquired the data. AAN and CM wrote the manuscript. AAN and CM and MJ, LM, CMM, IM provided methodological input and contributed to writing the manuscript. SN and TN edited and reviewed content of the manuscript.

## Ethics Declarations

Expedited ethical review approval of the study was obtained from the Rwanda National Ethics Committee (RNEC-IRB00001497). The Laboratory validation relied on retrospective data and biological samples which were put together as part of routine care of COVID-19 patients. Confidentiality has been respected through the use of codes instead of names.

## Consent for Publication

Not applicable.

## Competing Interests

The authors declare that they have no competing interests.

## Notes

### Competing Interest Statement

The authors have declared no competing interest.

### Author Declarations

Expedited ethical review approval of the study was obtained from the Rwanda National Ethics Committee (RNEC- IRB00001497). The Laboratory validation relied on retrospective data and biological samples which were put together as part of routine care of COVID-19 patients. Confidentiality has been respected through the use of codes instead of names. IRB review and oversight is not required because the activities involve secondary de-identified biospecimans.

